# How can the public health impact of vaccination be estimated?

**DOI:** 10.1101/2021.01.08.21249378

**Authors:** Susy Echeverria Londono, Xiang Li, Jaspreet Toor, Margaret J. de Villiers, Shevanthi Nayagam, Timothy B. Hallett, Kaja Abbas, Mark Jit, Petra Klepac, Kévin Jean, Tini Garske, Neil M. Ferguson, Katy A. M. Gaythorpe

## Abstract

Deaths due to vaccine preventable diseases cause a notable proportion of mortality worldwide. To quantify the importance of vaccination, it is necessary to estimate the burden averted through vaccination. The Vaccine Impact Modelling Consortium (VIMC) was established to estimate the health impact of vaccination. We describe the methods implemented by the VIMC to estimate impact by calendar year, birth year and year of vaccination (YoV). The calendar and birth year methods estimate impact in a particular year and over the lifetime of a particular birth cohort, respectively. The YoV method estimates the impact of a particular year’s vaccination activities through the use of impact ratios which have no stratification and stratification by activity type and/or birth cohort. Furthermore, we detail an impact extrapolation (IE) method for use between coverage scenarios. We compare the methods, focusing on YoV for hepatitis B, measles and yellow fever. We find that the YoV methods estimate similar impact with routine vaccinations but have greater yearly variation when campaigns occur with the birth cohort stratification. The IE performs well for the YoV methods, providing a time-efficient mechanism for updates to impact estimates. These methods provide a robust set of approaches to quantify vaccination impact.

## Introduction

Vaccination is one of the most effective interventions against infectious diseases and is estimated to prevent 2-3 million deaths annually, with an additional 1.5 million deaths could be averted with improvements in global vaccination coverage^1^. Vaccines can provide cost-effective, long-term protection and have resulted in the eradication of two major pathogens, rinderpest and smallpox, as well as the local elimination of others, such as measles^2,3^. Most vaccines have been shown to be safer than therapeutic medicines and are deemed second only to safe drinking water in terms of public health benefit^4–6^.

Of the burden that does occur due to vaccine preventable diseases, the majority is in Low and Middle Income Countries (LMICs)^7^. In many countries, the lack of reliable surveillance data and the inability to observe the burden that would have occurred without vaccination makes it difficult to directly calculate the mortality and morbidity prevented by vaccination. Hence, mathematical models are crucial for providing estimates of the burden averted by immunisation as they can project a no vaccination (counterfactual) scenario and scenarios with vaccination^8^. The Vaccine Impact Modelling Consortium (VIMC)^9^ was established in 2016, formally bringing together several modelling groups and a secretariat with a history of working together to estimate the impact of vaccines against ten pathogens, namely, Haemophilus influenzae type b (Hib), hepatitis B (HepB), human papillomavirus (HPV), Japanese encephalitis, measles, Neisseria meningitidis serogroup A (Meningitis A), rotavirus, rubella, Streptococcus pneumoniae and yellow fever (YF). Estimating the impact of vaccination is important as this reveals the effectiveness of current global vaccination strategies and whether any changes are needed.

Calculating vaccine impact is complex as for some pathogens, such as HepB and HPV, the consequences due to disease are not seen until long after infection. Thus, any vaccine impact is not immediately evident, but is still substantial leading to questions around how to appropriately attribute the impact of vaccination activities in order to inform the public and policy makers of their value. Vaccination activities also face a paradox as “vaccines are victims of their own success”^10^. For example, when the transmission of a fatal disease from an outbreak is prevented by vaccination, political and public support is assured. However, when vaccination is successful and the formerly-feared disease starts disappearing, the benefits of vaccination become less clear whilst the costs remain visible. As a consequence, public and political support may begin to decline. The benefits of the success of vaccination and control of diseases become less obvious with time, no longer being viewed with the same sense of urgency.

Through mathematical modelling of vaccination scenarios, we can explore the impact of vaccination activities stratified by characteristics such as birth cohort. Mathematical modelling, such as that carried out by the VIMC, is able to provide estimates of disease burden in terms of the number of cases, deaths and/or disability adjusted life years (DALYs). DALYs measure the years of healthy life lost due to disability from the disease and premature death. Comparing burden estimates in scenarios with and without vaccination can then quantify the burden averted by vaccination. However, impact may be attributed in different ways, some of which may be more appropriate in some settings than others. For example, one may be interested in the effect of vaccination on a particular birth cohort, so the impact of vaccination activities over the lifespan of that cohort needs to be aggregated. In contrast, for advocacy, planning and financing, it is valuable to ascertain the impact attributable to a particular year’s vaccination activities.

The VIMC uses vaccine coverage estimates from the World Health Organization (WHO), United Nations Children’s Fund (UNICEF) and Gavi, the Vaccine Alliance^11–13^. Due to changes and uncertainties in data on vaccination coverage, estimates of coverage are constantly refined and updated^12^, requiring estimates of vaccine impact to be recalculated accordingly. Whilst mathematical modelling of vaccine impact is vital to the process of the VIMC and ascertaining the effect of vaccination, it is not always feasible to conduct full model runs each time coverage is updated. As such, the VIMC secretariat has developed an impact extrapolation (IE) method whereby the impact ratio (impact per fully vaccinated person) from one modelling exercise can be applied to a new coverage scenario in order to extrapolate the impact calculation to the latest coverage estimates.

In this paper, we describe the methods that have been implemented by the VIMC to calculate the vaccination impact by calendar year, birth year and year of vaccination (YoV). To show how these methods perform, we use examples from HepB, measles and YF. Further, we describe an IE method which facilitates the estimation of impact from new vaccination coverage scenarios without full model runs. This is an approximation of the model projections which, whilst informative, is not a replacement for full model estimation. The IE does, however, allow for new approximate estimates to be produced very quickly and we detail a number of approaches each with their strengths and weaknesses.

## Methods

We detail a range of methods for calculating the impact of vaccination (*D*) defined as the burden averted for a given disease, country, year and, in some cases, birth cohort. Each method is used to address a specific point of interest, such as the number of lives saved by vaccination in a particular year (calendar year method), the number of individuals born in a particular year that will be saved due to vaccination (birth year method), and the number of individuals that will be saved due to a particular year’s vaccination activities (YoV methods). These usually involve the comparison of a focal scenario, the vaccination scenario we wish to assess, to a counterfactual or baseline scenario, which is the vaccination scenario we wish to compare against. We also detail the IE process and the scenarios carried out. All parameters and variables are defined in Table 1. A summary of the impact methods is shown in Table 3 and the impact ratios in Table 2.

**Table 1.** Parameter and variable definitions

| Parameter | Definition | Variable | Definition |
| --- | --- | --- | --- |
| $y$ | Year | $B_b$ | Burden in baseline/counterfactual scenario |
| $Y_v$ | Years of vaccination | $B_f$ | Burden in focal scenario |
| $Y_m$ | Years evaluated | $D_{b-f}$ | Difference or impact between baseline and focal scenarios ($B_b - B_f$) |
| $a$ | Age | $FVP$ | The number of fully vaccinated persons |
| $A_v$ | Age at routine vaccination | $\rho$ | Impact ratio (impact per FVP) |
| $A_m$ | Ages modelled | $D$ | Vaccine impact |
| $c$ | Country | | |
| $v$ | Vaccine | | |
| $k$ | Birth cohort | | |

**Table 2.**
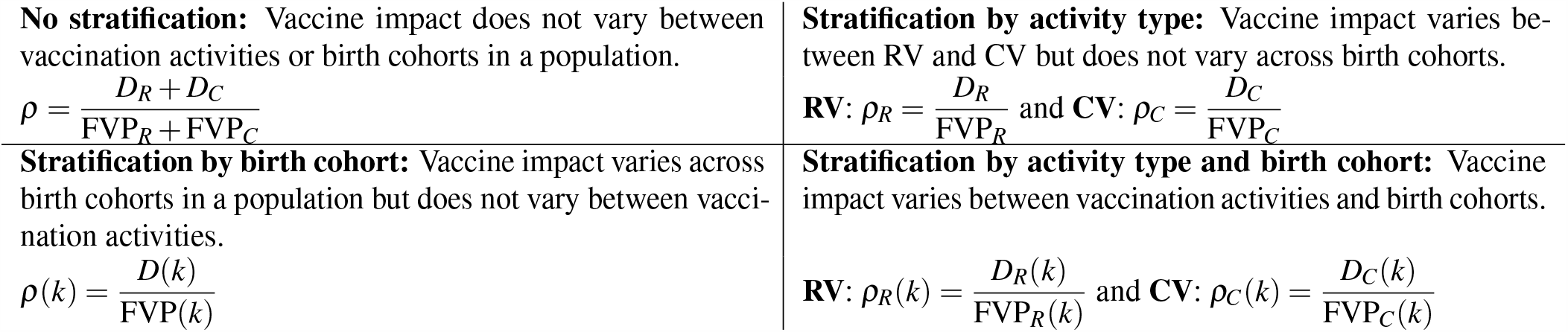
Stratifications of the impact ratios (*ρ*). Here, FVP_*R*_ and FVP_*C*_ denote fully vaccinated persons (FVPs) due to routine (RV) or campaign vaccination activities (CV) only; *D*_*R*_ and *D*_*C*_ denote impact due to RV or CV only, *D* denotes impact from both routine and campaign vaccinations, and *k* denotes a particular birth cohort.

**Table 3.** Summary of methods used by the VIMC for calculating the impact of vaccination (refer to Tables 1–2 for parameter values and for stratifications of the year of vaccination method, respectively).

| Method | Definition | Advantages | Limitations |
| --- | --- | --- | --- |
| Calendar year | Impact for a particular year | Intuitive. | Does not account for the long-term impact of vaccination on individual disease risk and cannot be linked to specific vaccination activities. |
| Birth year | Impact for a particular birth cohort | Captures the long-term effects of vaccination in protecting those vaccinated. | Duration of modelling needs to be pathogen-appropriate, e.g. the effects of HepB require a longer modelling time frame. Does not account for inter-cohort effects (e.g. herd protection). |
| Year of vaccination (YoV) | Impact attributed to year in which vaccination occurred | Possible to assess the effects of a specific year's activities and thus provide a direct comparison of the number of doses provided and their effect. This is useful for planning and economic purposes. | An impact ratio is required which can be affected by the years/activities or birth cohorts included, see Table 2 for details. Does not account for inter-cohort effects (e.g. herd protection). |

### Impact by calendar year (cross-sectional impact)

The calendar year method calculates the impact accrued over all ages for a specific year. It is generally the most intuitive and frequently used method to calculate the impact of vaccination. In this case, the vaccine impact, *D*_*b*−*f*_ (*a, c, y*), for age *a*, country *c* in year *y* is defined as the difference in disease burden between baseline and focal scenarios for a given year, *D*_*b*− *f*_ (*a, c, y*) = *B*_*b*_(*a, c, y*) −*B*_*f*_ (*a, c, y*). Here, the baseline scenario can have no vaccination or different coverage to the focal scenario. Aggregating the impact over all ages modelled (*A*_*m*_) means the impact for a country *c* in year *y* is given by 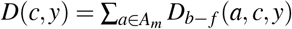.This does not account for the future disease burden averted through current vaccine activities.

### Impact by birth year (lifetime impact)

The birth year method accounts for the long-term impact accrued over the lifetime of a particular birth cohort *k*. For country *c*, this is given by *D*(*c, k*) = ∑_*y*−*a*=*k*_ *D*_*b*− *f*_ (*a, c, y*), where *y* ∈*Y*_*m*_ and *a* ∈*A*_*m*_. The duration of modelling needs to be appropriate to the pathogen of interest as in some cases, such as HepB, disease occurs later in life. For example, if we model vaccination for birth cohorts born from 2000 to 2030 and model disease burden until 2100, we do not account for the vaccine impact for those born in 2030 once they are over 70 years old. The method also does not specifically account for the impact of vaccinating a cohort outside the cohort vaccinated (e.g. because of herd protection).

### Impact by year of vaccination

The YoV methods are vital for determining the long-term impact of vaccination due to activities carried out in a particular year. We obtain the impact ratio, *ρ*, as the impact attributable per fully vaccinated person (FVP) calculated as the coverage × cohort size. This ratio can be stratified by different characteristics, such as birth cohort in order to catch temporal changes in transmission or healthcare or by activity type to capture the differing effects of routine and campaign vaccination (RV and CV, respectively); see Table 2 for details. The impact ratio allows effects due to a particular year’s worth of vaccination to be attributed to that year. Additionally, the impact ratio can then be projected with the IE method by updating the impact with a new estimate of FVPs; see impact extrapolation section for further details.

### Impact by year of vaccination: unstratified impact ratio

The simplest approach to calculate the impact by YoV is with an unstratified impact ratio which assumes that the effect of a vaccine is invariant over time and activity type. As such, the impact ratio, *ρ*, for a country, *c*, is the attributable vaccine impact divided by the number of FVPs:

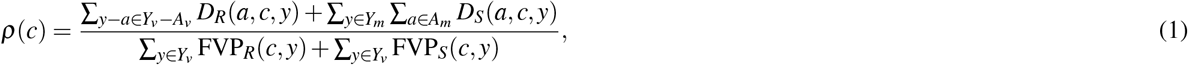

where *Y*_*v*_ − *A*_*v*_ refers to cohorts vaccinated in *Y*_*v*_ through RV.

Thus, the impact attributed to a year of vaccination is given by:

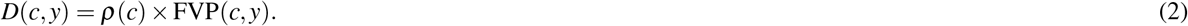

This method does not take into account any changes in treatment or transmission over time, nor the differing effects of vaccination through RV or CV.

### Impact by year of vaccination: impact ratio stratified by activity type

To calculate the impact by YoV using impact ratios stratified by activity type, we assume that RV and CV, which target multiple age groups, have different effects; for example due to dosage clustering. Hence, this method produces multiple, activity-specific impact ratios which can then be multiplied by the number of FVPs to calculate impact.

For RV, the impact ratio is defined as the impact for all cohorts who are vaccinated over time period *Y*_*v*_ per the additional FVPs between the baseline and focal scenarios. The impact for RI, *D*_*R*_, is given by 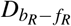, where *b*_*R*_ and *f*_*R*_ are the baseline and focal RI scenarios, respectively. Here, the impact ratio for RV is:

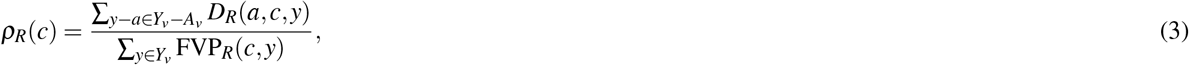

where *Y*_*v*_ − *A*_*v*_ are cohorts receiving vaccinations in years *Y*_*v*_. The impact by year of vaccination is then:

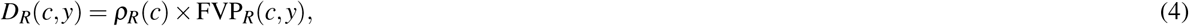

where FVP_*R*_ are FVPs vaccinated through RV. This does not account for the impact generated by indirect effects from and to cohorts not eligible for routine vaccination in *Y*_*v*_, nor does it account for interactions between RV and CV that may occur during or before years *Y* − *v*.

For CV, the impact ratio is averaged evenly over all ages across the entire time period (*Y*_*m*_). This is because we do not attempt to predict future CV coverage after the final year of credible campaign schedules. Therefore, the only impact due to CV comes from CV years *Y*_*v*_ and all campaign impact for birth cohorts born after this period can be attributed back to these vaccination years. The impact of CV, *D*_*C*_, is given by 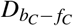, where *b*_*C*_ and *f*_*C*_ are the baseline and focal CV scenarios, respectively. The impact ratio is given by the following:

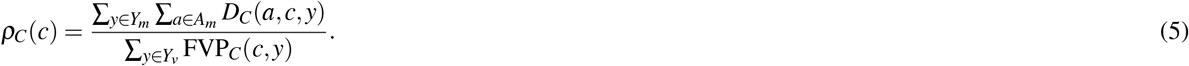

The impact by year of vaccination is then given by the following:

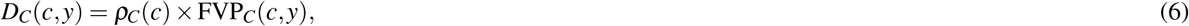

where FVP_*C*_ are FVPs vaccinated through CV.

Again, this does not account for interactions with the level of routine coverage across or before the same years.

The aggregated impact by YoV for both activities is the sum of the impact from RV and CV, i.e. sum of equations 4 and 6.

### Impact by year of vaccination: impact ratio stratified by birth cohort

In this method the impact ratio is invariant to vaccination activity type. However, vaccine effect is assumed to vary over time through birth cohorts. This means that rather than averaging the effect of vaccination over time, we account for the variation in transmission and health of the population. This influences how one year’s vaccination may work compared to another. For example, if therapeutic treatments for a disease improve over time, we may expect the impact of vaccination in 2050 to be less than that now as the population is generally healthier. In this case, the impact ratio is cohort specific, given by the following equations:

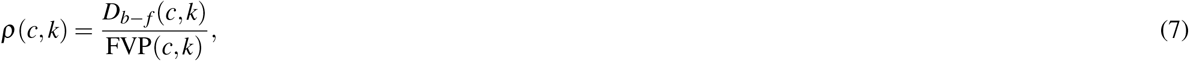

where *k* is the cohort defined by *k* = *y* − *a* and *a* ∈ *A*_*m*_. In order to find the impact attributed to one year of vaccination, all cohort specific impacts for those cohorts vaccinated in the year of interest must be aggregated. The impact by YoV is then given as follows:

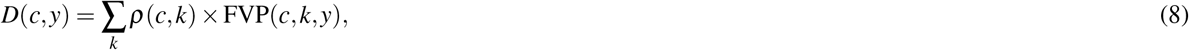

### Impact by year of vaccination: impact ratio stratified by both activity type and birth cohort

In the above methods, we illustrate the potential ways the impact ratio could change between birth cohorts or by vaccination activity. However, it is likely that in reality the impact varies with respect to both of these aspects. In the following method, we use the approach of the activity stratification over birth cohort to arrive at an impact ratio per birth cohort *and* activity. The impact ratios by activity (R for RV; C for CV) are thus the following:

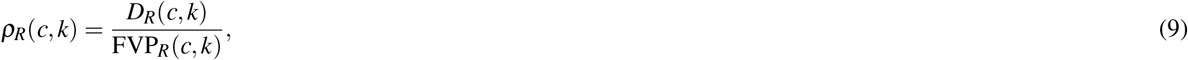

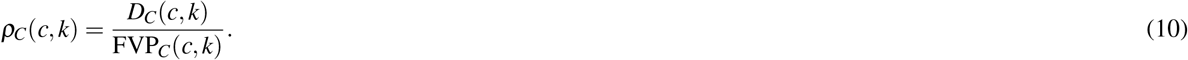

Therefore the impact by YoV is provided by the sum of the activity and cohort-specific components:

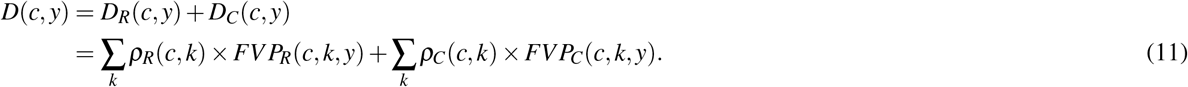

Note that in the above method the numerator may also be affected by vaccination activities experienced by cohorts around them, so not captured in the denominator. Hence the impact ratio could potentially be infinite.

## Summary of impact methods

We summarise the key features of each vaccine impact method in Table 3.

### Impact extrapolation

As coverage estimates are frequently updated within the WHO/UNICEF Estimates of National Immunization Coverage (WUENIC) and Gavi’s Operational Forecast (OP)^11–13^, one of the vital VIMC tasks is to provide updated impact estimates to inform future RV and CV. Translating these coverage estimates into tangible estimates of vaccine impact in terms of mortality and morbidity averted is crucial. However, given the regular updates of vaccination coverage and due to the effort and time required for full model updates across the VIMC, a simplified process, the IE, has been developed to update past rounds of impact estimates with new coverage data.

The IE is primarily a linear interpolation of current vaccine impact estimates with new coverage estimates assuming constant country- and delivery strategy specific rates of mortality and morbidity averted per dose of vaccine. It can be applied to the YoV methods as these provide us with an impact ratio (*ρ* in Table 2). Hence, when coverage is updated, the updated number of FVPs can be calculated by multiplying the new coverage with the target population. The updated FVPs (FVP^∗^) are then multiplied by the previously calculated *ρ* to calculate the updated impact (*D*^∗^):

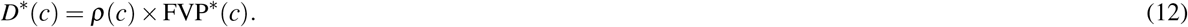

### Models and vaccination scenarios

As the burden in each vaccination scenario is calculated using mathematical models of pathogen transmission dynamics and implementation of vaccination activities, the impact of vaccination will vary depending on the underlying model being used. The models within the VIMC vary from being static, whereby the direct effects of vaccination on FVPs are modelled assuming that pathogen transmission intensity is not modified by vaccination coverage, to dynamic, whereby infectious disease transmission dynamics, direct effects of vaccination on FVPs and herd effects are modelled. Herd effects account for any indirect effects of immunity (both natural and/or from vaccination activities) due to a reduction in transmission. For example, an increase in FVPs offers indirect protection as it reduces the risk of a non-vaccinated individual coming into contact with the disease. Herd effects will not arise for all vaccine preventable diseases, for example, Japanese Encephalitis is vector-borne (i.e. not transmitted person-to-person) and the majority of cases are due to spillover events^14^. Hence in this case, FVPs will not provide protection to others.

In this analysis, we focus on calculating impact estimates for HepB (dynamic model with long-term outcomes and herd effects), measles (dynamic model with short-term outcomes and herd effects) and YF (static model without herd effects)^15–18^. We use anonymised countries for each of the pathogens, denoted by Country A for HepB, Country B for measles and Country C for YF, with a focus on the years 2000–2017. Standardised demographic data (live births per year, death rates) are based on the 2017 United Nations World Population Prospects (UNWPP)^19^. Immunisation coverage from 1980 to 2016 corresponds to the WUENIC estimates of National Immunization Coverage as published in July 2017^12^, and 2017–2030 coverage corresponds to Gavi’s October 2017 Operational forecast (OP)^13, 15^. Using each of the vaccine impact methods, we calculate the number of deaths averted due to vaccination over the years 2000–2017. Though we only show this for 2000–2017, we are taking into account the impact of vaccination activities from 2000 to 2030.

We also investigate the performance of the IE under the YoV methods with no stratification, stratification by activity and cohort separately and in combination by applying it to HepB, measles and YF in Countries A, B and C with updated coverage data. More specifically, for each of these pathogens, we update RI coverage from 1980 to 2017 based on WUENIC estimates published in July 2018^12^, and 2018–2030 routine and campaign coverage is updated to Gavi’s October 2018 OP^13^. Changing coverage of activities alters the number of FVPs (population demography and models remain the same). We then re-run the models with this updated coverage scenario and compare to the IE projections in order to determine the reliability of the IE.

All of the analyses were carried out in R and the impact methods are available through the R package *vimpact* on github (https://github.com/vimc/vimpact)^20^.

## Results

### Estimates of the impact of vaccination

From 2000 to 2017, HepB, measles and YF have different vaccination activities in each of the countries. We analyse the impact calculation methods for RV alone and in combination with CV. We find that the estimated impact of vaccination varies depending on the method used.

There are only RV for HepB which are given as a birth dose (soon after birth) and as infant doses (<1 year old) (Fig.1A). As HepB-attributable mortality primarily affects those over 40 years of age, we find that the birth year method captures a greater impact as it accrues the long-term benefits of vaccination over the lifetime of birth cohorts. With the calendar year method accruing the impact for all ages over a particular year, this long-term impact is not captured (Fig.1B). Prior to 2013, YF has routine infant dose vaccinations (<1 year old) only with the calendar year method capturing slightly more impact (relative to HepB) as yellow fever largely affects those under 30 due to natural immunity acquired with age in older adults (Fig. 1A–1B). For measles from 2000–2017 in Country B, CV occur alongside RV. RV provides the first dose of a measles vaccine to those aged 1 or under (Fig.1A). As FVPs accumulate, the birth year and calendar year methods show an increasing number of deaths averted over time. In comparison to HepB where it is vital that the long-term benefits are captured, the calendar year method shows a high impact as measles-related mortality is focused in the under-5s (Fig.1B).

**Figure 1.**
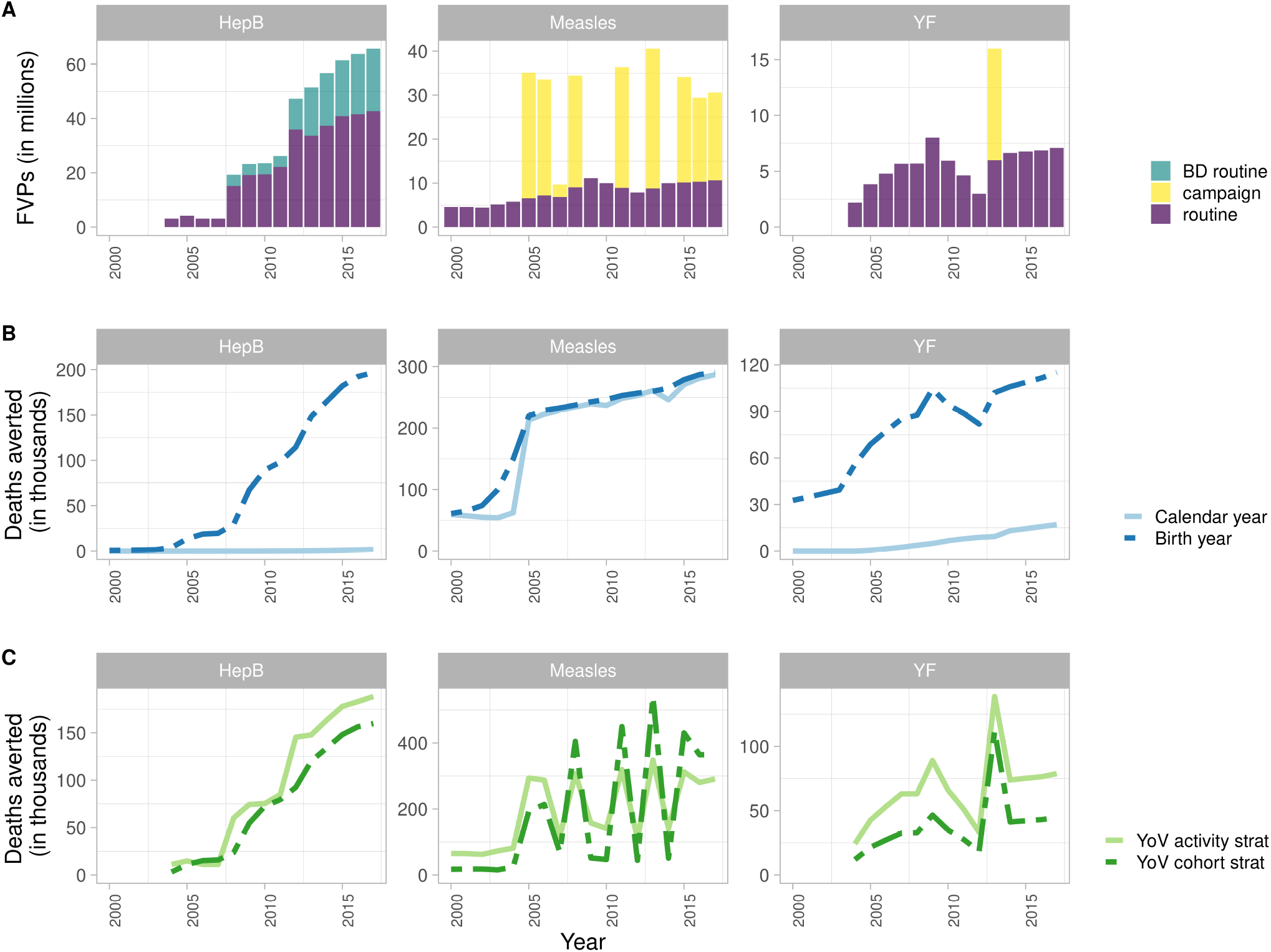
Fully vaccinated persons (FVPs) and mean estimates of deaths averted for Hepatitis B (HepB) in Country A, measles in Country B and yellow fever (YF) in Country C from 2000 to 2017. (A) FVPs for HepB birth dose (BD routine) and infant dose (routine) routine vaccination activities. FVPs for first routine dose of a measles containing vaccine and measles campaign activities. FVPs for YF routine and campaign activities. (B) Impact by calendar and birth year methods. (C) Impact by year of vaccination (YoV) with impact ratio stratified by activity type and birth cohort.

The impact of vaccination estimated by the YoV methods varies over time with the activity stratification showing an averaged impact and the birth cohort stratification capturing gradual improvements in population health, particularly when herd effects are modelled. Hence, the activity stratification initially shows a higher number of deaths averted than the birth cohort stratification. Accordingly with RV only for HepB and prior to 2013 for YF, we see that the impact calculated by the activity stratification is greater as it is averaging the impact over all FVPs. The impact for the birth cohort stratification does not surpass the activity stratification within the time frame of this analysis for these two pathogens (Fig.1C). Notably for HepB, with RV occurring in the first year of life, the impact by year of vaccination and impact by birth year methods show a similar impact.

For YF, there is one CV in 2013 which results in a peak in the deaths averted with the largest impact seen with the activity stratification. The birth cohort stratification shows a lower impact as this is the first campaign (Fig.1C). For measles, we see the same pattern for the first campaign but for subsequent campaigns we see a higher impact with the birth cohort stratification because it is capturing the overall improvement in population health. Furthermore, herd effects are modelled for measles which contribute to the birth cohort stratification impact increasing as herd effects builds up reducing transmission in the population (Fig.1C).

### Performance of the impact extrapolation method with updated coverage compared to model estimates

We explore the IE under the YoV methods with impact ratios stratified as per Table 2 for scenarios where vaccination coverage is updated from 2017 estimates to those published in 2018 (as described in Methods; change in FVPs shown in Figures 2A and 3A). The accuracy of the IE projections is determined by comparing to model estimates given the same updated coverage. Vaccine impact, i.e. deaths averted, by year of vaccination and by year of birth are visualised in Fig. 2 and Fig. 3, respectively, for Countries A, B and C over the years 2000–2017. The total relative difference in deaths averted over 2000–2017 between the IE and model estimates are shown in Table 4. We find that the accuracy of the IE varies depending on the impact ratio stratification method, underlying model dynamics and pathogen characteristics.

**Table 4.** Relative total difference of impact extrapolation (IE) to model estimates for deaths averted over 2000–2017 (%) by year of vaccination (YoV) and year of birth (YoB) due to vaccination activities for hepatitis B (HepB) in Country A, measles in Country B and yellow fever (YF) in Country C. Impact estimated by the year of vaccination stratification methods as shown in Figures 2-3. Negative numbers correspond to the IE underestimating and positive numbers correspond to the IE overestimating the number of deaths averted over 2000–2017.

| Disease and deaths averted by YoV/YoB | No stratification | Activity type stratification | Birth cohort stratification | Activity type and birth cohort stratification |
| --- | --- | --- | --- | --- |
| HepB- YoV | -0.86 | 7.24 | 0.40 | 0.30 |
| HepB- YoB | -4.12 | -0.35 | 0.40 | 0.04 |
| Measles- YoV | -8.90 | -0.76 | -0.02 | -0.81 |
| Measles- YoB | -0.51 | -4.04 | -4.34 | -4.10 |
| YF- YoV | -12.16 | -0.84 | -8.80 | -0.67 |
| YF- YoB | -10.37 | -7.41 | -8.62 | -4.66 |

**Figure 2.**
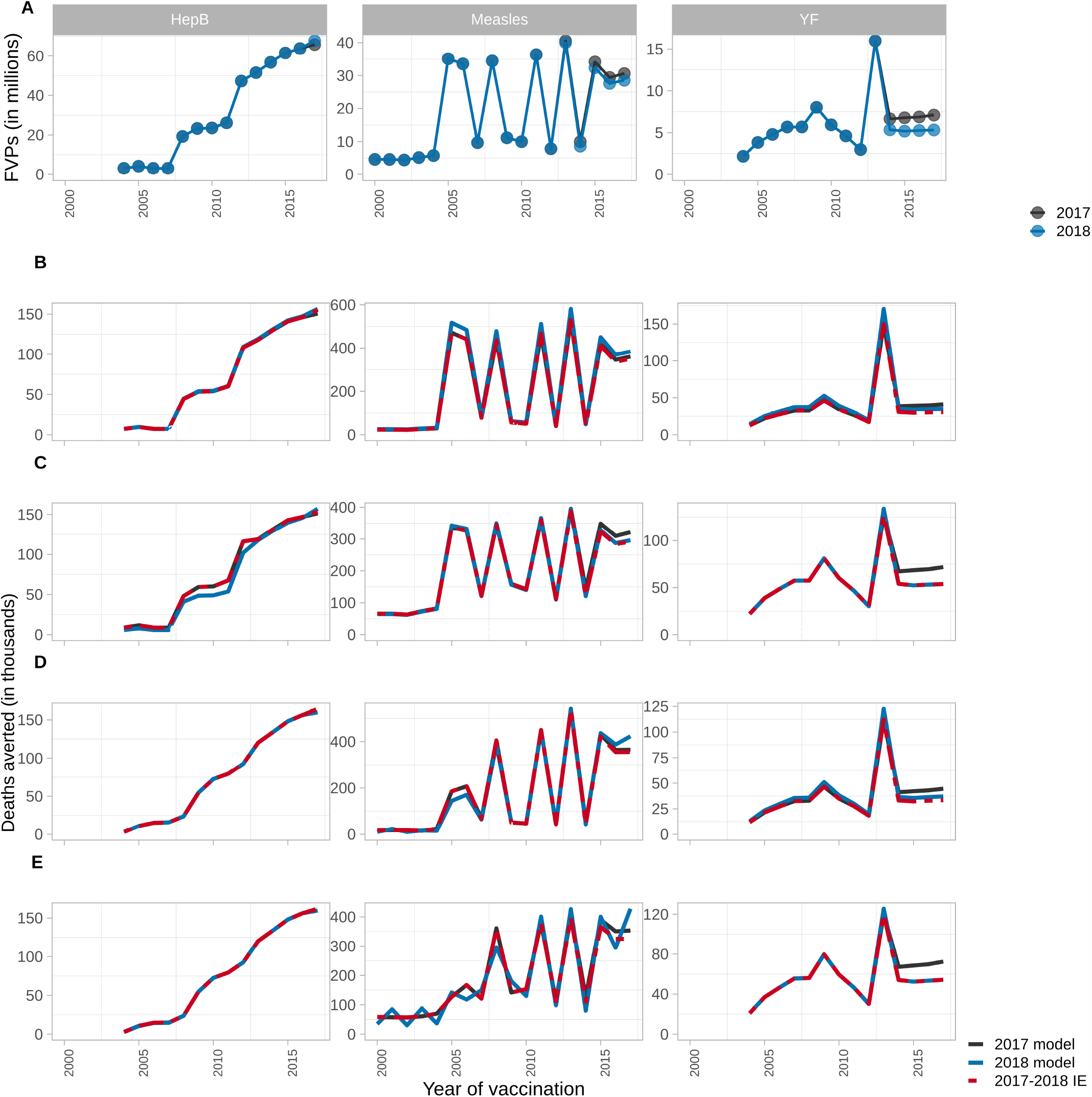
Model estimates and impact extrapolation showing deaths averted by year of vaccination per year for hepatitis B (HepB) in Country A, measles in Country B and yellow fever (YF) in Country C from 2000 to 2017 using 2017 coverage (2017 model), 2018 coverage with corresponding model estimates (2018 model), and 2018 coverage with the impact extrapolation (2017–2018 IE). (A) Fully vaccinated persons (FVPs) in 2017 and 2018. (B) Impact by year of vaccination with unstratified impact ratio. (C) Impact by year of vaccination with impact ratio stratified by activity type. (D) Impact by year of vaccination with impact ratio stratified by birth cohort. (E) Impact by year of vaccination with impact ratio stratified by activity type and birth cohort.

**Figure 3.**
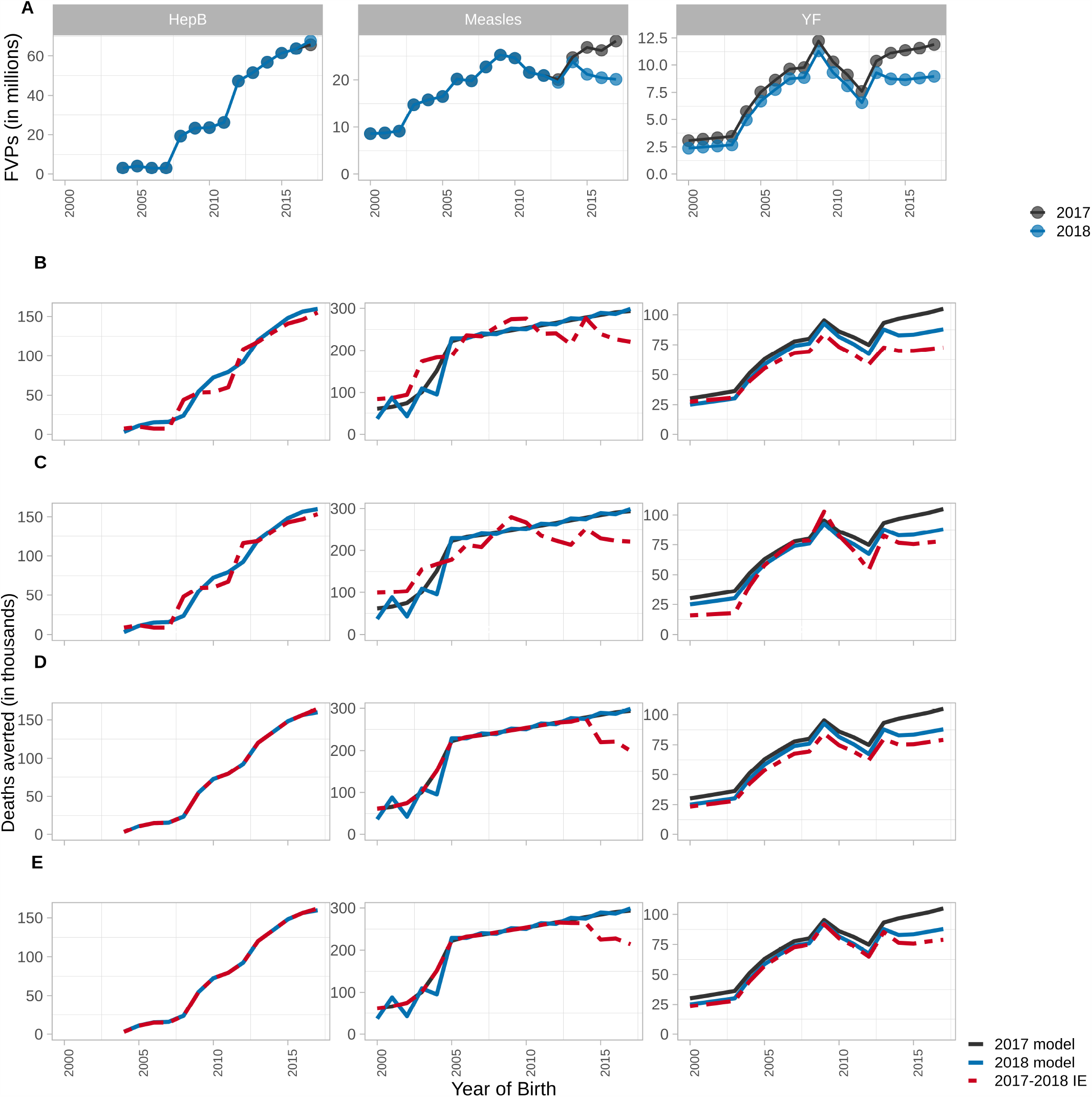
Model estimates and impact extrapolation showing deaths averted by year of vaccination per birth year for hepatitis B (HepB) in Country A, measles in Country B and yellow fever (YF) in Country C from 2000 to 2017 using 2017 coverage (2017 model), 2018 coverage with corresponding model estimates (2018 model), and 2018 coverage with the impact extrapolation (2017–2018 IE). (A) Fully vaccinated persons (FVPs) in 2017 and 2018. (B) Impact by year of birth with unstratified impact ratio. (C) Impact by year of birth with impact ratio stratified by activity type. (D) Impact by year of birth with impact ratio stratified by birth cohort. (E) Impact by year of birth with impact ratio stratified by activity type and birth cohort.

Using impact ratios without stratification, the IE performs relatively well for HepB in comparison to measles and YF (Table 4). As HepB only has RV, the activity type does not need to be taken into account. Furthermore, as this is given as a birth or infant dose, unstratified impact ratios would not capture variation in effect but this change would likely occur subtly over long time periods. Both RV and CV occur for measles and YF for which the IE with unstratified impact ratios underestimates the deaths averted due to the drop in coverage/FVPs in the 2018 coverage scenario prior to 2020 (Figs. 2A,B and 3A,B).

The IE with impact ratios stratified by activity does not perform well for HepB relative to measles and YF as it overestimates the impact associated with a change in coverage (Table 4; Figs. 2C and 3C). In the updated coverage scenario, HepB coverage improves over the years 2017–2022 creating more FVPs. With the activity stratification, this coverage improvement is being averaged over the whole time period. As such, earlier RV are artificially inflated. Additionally, as HepB-attributed disease occurs later in life, vaccinating more people born in later years will produce herd protection for those born over 2000–2017.

For measles, the activity stratification viewed by year of vaccination performs well (Table 4; Figs. 2C and 3C). Similar to HepB, measles coverage for RV improves greatly post-2020 with the 2018 coverage scenario. However, the IE does not overestimate the impact of this in the same way as it did for HepB because measles is focused in the under-5s so creating more FVPs later on does not produce herd effects for the 2000–2017 birth cohorts.

Using the activity stratification for YF, with changes to coverage, the IE works well with only a small underestimation of deaths averted (0.84%) by year of vaccination over 2000–2017 compared to model estimates (Fig. 2C) as it is a static model with no herd protection. However, there is larger underestimation of deaths averted (7.41%) by year of birth (Fig. 3C) as the FVPs vaccinated in the 2000–2017 birth cohorts is lower in the 2018 coverage scenario (Fig. 3A). Furthermore, compared to the unstratified impact ratio, the impact ratio for CV specifically is smaller and more sensitive to a reduction in FVPs. In contrast, the impact ratio for RV is larger than unstratified which accounts for the fact that RV will almost always be given to unvaccinated infants, as opposed to CV which, as dose distribution is assumed to be random, may revaccinate individuals who are already immune.

Using the birth cohort stratification, the IE performs well for HepB but underestimates the number of deaths averted by year of vaccination and birth year under the updated coverage scenarios for measles and YF (Table 4; Figs. 2D and 3D). Although the 2018 coverage scenarios generally have higher coverage post-2020, the coverage drops prior to 2020 for measles and YF but for HepB it improves slightly (Figs. 2A and 3A). As the birth cohort stratification captures greater yearly variation, it does well for the smaller change in HepB coverage and shows a corresponding decline in impact for measles and YF as it is more sensitive to changes in coverage than the activity stratification. This captures a limitation of the birth cohort stratification as it assumes the impact from RV and CV will be similar when they could be distinctively affected by dose wastage i.e. a RV will almost always be given to a susceptible child whereas campaigns may vaccinate individuals who have already been vaccinated, thus non-intuitively having a smaller impact.

Implementing the IE when stratifying by both activity and birth cohort results in the IE being sensitive to a combination of factors that affect the activity and birth cohort stratifications, such as the type of activities occurring, underlying model dynamics and yearly coverage changes. For measles and YF, this stratification captures a decline in coverage prior to 2020 which does not occur for HepB. Additionally it captures any herd protection arising from post-2020 coverage improvements to varying extents (seen more for HepB than measles because of differing age dynamics; Table 4; Figs. 2E and 3E).

When determining the accuracy of the IE, it is useful to look at both the relative total difference in deaths averted over 2000–2017 between the IE and model estimates (Table 4), and the difference across each year (Fig. 2 and Fig. 3). Some of the under- and over-estimation of the IE that is seen across the years can cancel out when looking at the aggregated difference over the whole time period. As such, the IE performs well when looking at the total difference for no stratification viewed by year of birth for measles, the activity stratification viewed by year of birth for HepB and the birth cohort stratification viewed by year of vaccination for measles (Table 4). However, when focusing on the difference in a specific year, the IE is seen to under- and over-estimate in certain years (Fig. 3B, Fig. 3C and Fig. 2D).

Overall, in our examples, we have shown that the IE works well and is an effective tool to update vaccine impacts. Notably, the activity stratification is accurate for static models with dose dependency but may overestimate the impact of coverage improvements when dynamic herd protection comes into play. The birth cohort stratification may underestimate impact if coverage declines before improving in later years.

Caution needs to be taken when deciding the appropriateness of implementing an IE. In cases where coverage is reduced more drastically or an activity is delayed, if the pathogen poses a risk of outbreaks and the model is dynamic, the IE may not be accurate as it may miss capturing the impact of any outbreaks that may occur during this period. With larger delays to activities, the risks of the IE not capturing such outbreaks would rise. Additionally, if a follow-up activity cannot reach the individuals missed, using the IE may not be accurate. In such cases, new model runs would be required to provide updated estimates of vaccine impact.

## Discussion

We have presented the main methods used by VIMC to estimate vaccine impact (summarised in Table 3). We also have shown how these methods perform when providing an IE. Each of the methods differ and the appropriate method will depend on the perspective of interest.

The vaccine impact methods primarily differ by how they attribute impact. The calendar year method attributes impact to the year examined irrespective of which age groups are targeted and when the vaccination occurred. In contrast, impact by birth year attributes impact to the birth cohorts of interest, irrespective of when the vaccination occurred over their lifespan. When only RIs occur in the first year of life, the impact by YoV and impact by birth year will be similar. However, when vaccination activities vary in who is targeted and when, the estimation of impact becomes complex.

The two impact by YoV methods differ not in *when* they attribute impact but in the assumptions around how the effect of vaccination may change over time or by delivery method. The birth cohort stratification accounts for changes in transmission and healthcare over time, allowing the impact of vaccination to be adjusted given improvements in other interventions such as treatment or the effects of climate change on vector-borne diseases as seen for YF^21,22^. The issue with this method is that it may be sensitive to the introduction of RIs or SIAs as these dictate the vaccination experience of cohorts to come, for example, a birth cohort born the year after a mass vaccination campaign should be well protected by the population vaccinated in that campaign. This characteristic is a strength in that it accounts for herd effects. However, the denominator for this method does not include vaccinations given to birth cohorts vaccinated outside the year of interest that might have an indirect (herd) effect on the birth cohort of interest. Furthermore, as the vaccination coverage, implementation and, arguably, demography are all subject to high degrees of uncertainty, this characteristic will also exacerbate any inaccuracies.

The most accurate way to capture herd effects would be to repeatedly run the underlying mathematical models incrementally, i.e. starting with the baseline and adding vaccination of a single birth cohort, year and activity type with each iteration. This has the advantage of correctly attributing the full indirect impact of each additional set of vaccinations to the whole population. The disadvantage of this method is that it is computationally expensive.

For computational and time efficiency, the IE is used by the VIMC to approximate the impact of vaccination given a new scenario around FVPs relative to the one modelled. This means that uncertainties in the modelling process are propagated into the IE. The update was developed in order to provide estimates on a short timescale given the latest available demographic and vaccination data. We show that impact by YoV methods perform well in many situations and given updates in coverage, each stratification would be appropriate for their related insights. However, there are limitations to the IE, particularly for diseases that pose a risk of outbreaks and that are modelled dynamically, if there are drastic changes to activities (e.g. coverage drops or delays to activities) as the IE may not account for rises in case numbers during such periods.

The methods shown are not a complete set, and as further research questions develop, new methods for calculating vaccine impact will need to be incorporated. Though the YoV methods capture changes over time due to healthcare or transmission variation and the different effects of vaccination activity, they do not capture issues around vaccination and healthcare clustering which has been shown to be influential^3,23^. An extension would be to account for the potential of re-vaccinating the same individuals each time, a method that could highlight and identify groups of zero-dose children and health equity^24^. Similarly, the current activity stratification allows us to further explore questions around vaccine delivery and health access. Finally, the results shown are presented nationally, whereas RIs and SIAs may be delivered sub-nationally, as well as the disease transmission and health access being spatially heterogeneous especially for large countries. As such, the next steps will be to examine the sensitivity to these national assumptions and account for the spatial heterogeneity in vaccine impact.

Whilst vaccination is one of the most effective interventions against infectious diseases, the specific implementation of vaccination requires substantial planning, support and financing. Calculating the public health impact of vaccination and understanding the different methods for doing so is vital. We have presented multiple methods to express the complex effects of vaccination which will continue to be developed for ever increasing questions around optimal vaccine use.

## Data Availability

Data available on request but will be available through an online data visualisation tool accessible through vaccineimpact.org, and attached to the forthcoming paper: https://www.medrxiv.org/content/10.1101/19004358v1.
Code is available at: https://github.com/vimc/vimpact

## Author contributions statement

XL, TG, NMF developed the methods; SEL, XL, JT, KAMG, TG conducted the analyses; SEL, XL, JT, KAMG, TG analysed the results; JT, SEL, XL, KAMG wrote the first draft; MJdV, SN, TBH modelled Hepatitis B; KA, MJ modelled measles; TG, KAMG, KJ modelled yellow fever. All authors edited and reviewed the manuscript.

